# Same zone diameter, different verdict: clinical and surveillance implications of EUCAST 2025 versus CLSI 2025 breakpoint discordance among Enterobacterales, *Staphylococcus* and *Pseudomonas* isolates at Buea Regional Hospital, Cameroon

**DOI:** 10.64898/2026.10.20.26366059

**Authors:** Kukwah Anthony Tufon, Henry Dilonga Meriki, Kamsang Kingsly Ngong, Ayah Flora Bolimo, Youmbi Sylvain Kouanou, Nkai Gideon Nyenty, Bsadjo Tchamba Gertrude, Lie Banlock Guymax Legrand, Bassuke Blair Ewane, Frednah Moiwo Tutuwan, Eyong Mathias Esum

## Abstract

**Background:** AST guide both patient management and national AMR surveillance. In Cameroon, laboratories interpret AST using either CLSI or EUCAST standard which may not always give the same conclusion for every case. This study compared EUCAST 2025 and CLSI 2025 interpretations of identical disk diffusion results at Buea Regional Hospital (BRH).

**Methods:** This was a retrospective laboratory-based study where raw inhibition zone diameters of Enterobacterales, *Staphylococcus* and *Pseudomonas* isolates recorded in the BRH WHONET database (2023 – 2025) were interpreted with EUCAST 2025 and CLSI 2025 standards. Agreement was evaluated using Cohen’s kappa (κ), with κ < 0.61 considered poor agreement. Major interpretation switches (susceptible ↔ resistant) were enumerated, and the standard more associated with non-susceptible outcome was identified using the exact McNemar test.

**Results:** A total of 1,198 isolates (748 Enterobacterales, 367 *Staphylococcus* and 83 *Pseudomonas*) were included in the analysis. Poor agreement was observed for 4/12 (33.3%) antibiotics against Enterobacterales (lowest: ofloxacin, κ = 0.48), 5/10 (50%) against *Staphylococcus* (lowest: levofloxacin, κ = 0.08) and 5/7 (71.4%) against *Pseudomonas* (lowest: cefepime and imipenem, κ = 0.24). Major interpretation switches occurred for 3/12 antibiotics (Amox/clavulanic acid [1.1%], ceftriaxone [0.6%] and ofloxacin [15.6%]) against Enterobacterales, 6/10 against *Staphylococcus* (minocycline [21.1%], gentamicin [15.9%], rifampicin [14.4%], clindamycin [2.7%], tetracycline [4.5%], levofloxacin [13.7%]) and 2/7 among *Pseudomonas* (cefepime [20%], ciprofloxacin [3%]). Overall, EUCAST was significantly more likely (p < 0.001) to classify isolates as non-susceptible.

**Conclusion:** Poor agreement between EUCAST 2025 and CLSI 2025 affected one third of the antibiotics tested for Enterobacterales, half for *Staphylococcus* and close to three quarters for *Pseudomonas*. Major interpretation switches were quite common for levofloxacin, ofloxacin, gentamicin, minocycline, rifampicin and cefepime. EUCAST was generally more likely to classify isolates as non-susceptible. A single national breakpoint standard is recommended in Cameroon so as to optimize the reliability of its surveillance data.

## Introduction

Antimicrobial resistance (AMR) is one of the top threats to global health with more than 39 million attributable deaths forecasted between 2025 and 2050 [1]. In 2023, one in six laboratory-confirmed bacterial infections worldwide was caused by antibiotic resistant bacteria. to antibiotic treatment [2]. The burden of bacterial AMR is high in World Health Organization (WHO) African region and laboratory capacity to measure it remains limited [3]. Because such estimates are assembled from routine laboratory reports, their validity rests on how each individual antimicrobial susceptibility result is generated and categorized.

Antimicrobial susceptibility testing (AST) supports both individual treatment decisions and AMR surveillance. In most laboratories across the world, raw inhibition zone diameters are converted into interpretive categories using clinical breakpoints obtained from either the European Committee on Antimicrobial Susceptibility Testing (EUCAST) or the Clinical and Laboratory Standards Institute (CLSI). Both standards derive breakpoints from microbiological, pharmacokinetic/pharmacodynamic (PK/PD) and clinical outcome data [4], yet they differ in breakpoint values and category definitions. Both bodies also revise their breakpoints periodically leading to new set cut-off values for some organism-antibiotic specific combination. Because the two standards do not always set their cut-off values at the same zone size, an isolate with a given inhibition zone diameter may be classified as susceptible under one standard but not under the other.

These differences in classification have practical consequences for both patient care and surveillance. The same measurement reported as susceptible under one standard and non-susceptible under the other may alter both the result issued for an individual patient and the AMR surveillance data derived from aggregated results [5]. A switch from CLSI to EUCAST in some hospitals brought about a shift in cumulative antibiograms and multidrug resistance rates for some organism–antibiotic combinations without any change in the underlying bacterial populations [6,7]. Comparative studies of the two standards have been conducted in some countries [8–10]. Most of these studies, however, compared older editions of the standards and used minimum inhibitory concentrations (MICs) from automated systems. Little is known about how the 2025 editions compare for disk diffusion, the predominant AST method in many African laboratories. In addition, this gap is most significant in settings where surveillance systems combine results interpreted under both standards.

Cameroon depicts this situation. AMR has been documented in human, animal and environmental isolates across the country [11], and resistance trends have been described [12]. National surveillance relies on sentinel laboratories that enter AST data into WHONET for transmission to the national level [13]. Laboratories contributing to the program may interpret AST results using either CLSI or EUCAST breakpoints, and their data are pooled into national statistics. To the best of our knowledge, the differences in disk diffusion zone diameter classification of the two standards have not been assessed in this setting even though laboratories are allowed to use either of the standards.

This study therefore compared CLSI 2025 and EUCAST 2025 interpretations of the same disk diffusion results among Enterobacterales, *Staphylococcus* and *Pseudomonas* isolates at Buea Regional Hospital, Cameroon. Specifically, it aimed to identify antibiotics with poor agreement between the two standards, identify antibiotics with major interpretation switches (susceptible↔ resistant) that could change clinical decisions, and to determine which standard is more likely to classify isolates as non-susceptible in the study area.

## Materials and methods

### Study site, design and population

This was a retrospective, laboratory-based comparative study carried out at Buea Regional Hospital (BRH), a public referral hospital in Buea, the administrative capital of the South West Region of Cameroon. BRH serves Buea Health District and neighbouring districts, receiving patients from different communities. Its microbiology laboratory performs routine bacterial culture, identification and AST by disk diffusion, records results in WHONET and contributes AST data to Cameroon’s national AMR surveillance program. The study used AST records of Enterobacterales, *Staphylococcus* and *Pseudomonas* isolates recovered from routine clinical specimens between January 2023 and December 2025. The period of data accession/collection was 5^th^ – 10^th^ May 2026. All eligible isolates within this period were included.

### Data collection and management

Data were extracted from the BRH WHONET database into Microsoft Excel (version 2021) by authorized laboratory personnel. The investigators worked only with the de-identified dataset. The variables used for this study included specimen type, year of isolation, organism identification, and raw inhibition zone diameters. Organism nomenclature across the data set was harmonized, specimen entries were verified, duplicate records were identified and one deleted. Entries with incomplete/inconsistent relevant laboratory records, organism–antibiotic results for which no applicable breakpoint existed in one of the two standards, or for which the two standards specify different disk contents were excluded from the study.

### Breakpoint interpretation

Raw measured inhibition zone diameters were interpreted independently using the EUCAST breakpoint tables version 15.0 (2025) [14] and the CLSI M100 35th edition (2025) [15]. For each isolate and antibiotic, the zone diameter column was retained and two interpretation columns were added, one for EUCAST 2025 and one for CLSI 2025. Under CLSI 2025, results were categorised as susceptible (S), intermediate (I), susceptible-dose dependent (SDD, where defined) or resistant (R) where applicable. Under EUCAST 2025, results were categorised as susceptible, standard dosing regimen (S), susceptible, increased exposure (I) or resistant (R) where applicable. Only organism–antibiotic combinations with zone diameter breakpoints in both standards and the same disk content were compared. Genus-specific, species-specific and site-specific cut offs for each standard were taken into consideration where applicable (for example, species-specific EUCAST breakpoints for *S. aureus* versus coagulase-negative staphylococci were applied where applicable). The choices and number of antibiotics tested for each group of organisms were based on what the two standards recommended.

### Ethical consideration

Ethical approval was gotten from the South West Regional Ethics Committee for Human Health Research (Ref. No. 1070/CRERSH/SW/C/04/2026). Administrative authorisation was obtained from the South West Regional Delegation of Public Health (Ref. No. 850/MINSANTE/SWR/RDPH/RCB.PT/04/2026) and from the administrative authorities of BRH. All patient identifiers (names, age, sex, laboratory numbers, addresses etc) were removed before the dataset was shared with the investigators, so individual informed consent was not mandatory.

### Data analysis

The data was analysed using IBM SPSS Statistics version 25. Frequencies and proportions described the distribution of isolates. Agreement between EUCAST 2025 and CLSI 2025 categories was assessed by percentage concordance (proportion of isolate–antibiotic results assigned the same category under both standards) and by Cohen’s kappa (κ) with 95% confidence intervals (CI). Kappa values were interpreted using the Landis and Koch scale

[16] where 0.61–0.80 and 0.81–1.00 are considered as substantial agreement and almost perfect agreement respectively. Antibiotics with κ < 0.61 were considered as poor agreement.

Category differences were classified as major interpretation switches (S ↔ R) or as exposure- or dosing-related switches (S ↔ I/SDD or I/SDD ↔ R), and their frequencies and proportions were calculated. Non-susceptible was defined as I, SDD or R. Exact McNemar test was used compare paired susceptible versus non-susceptible classifications under the two standards. Statistical significance was set at p < 0.05.

## Results

A total of 1,198 bacterial isolates were analysed in this study: 748 Enterobacterales, 367 *Staphylococcus* and 83 *Pseudomonas*. Of the 748 Enterobacterales, *Escherichia coli* (n = 313, 41.8%) and *Klebsiella pneumoniae* (n = 127, 17%) were the most frequently isolated (Table 1). Coagulase-positive *Staphylococcus* (n = 228, 62.1%) was the most frequently isolated *Staphylococcus*.

**Table 1.** Distribution of Enterobacterales, *Staphylococcus* and *Pseudomonas* isolates at Buea Regional Hospital, January 2023 to December 2025.

| Group | Organism | n | % within group |
| --- | --- | --- | --- |
| Enterobacterales ($n = 748$) | *Escherichia coli* | 313 | 41.8 |
|  | *Klebsiella pneumoniae* | 127 | 17.0 |
|  | *Enterobacter cloacae* | 71 | 9.5 |
|  | *Serratia liquefaciens* | 33 | 4.4 |
|  | *Proteus mirabilis* | 23 | 3.1 |
|  | *Serratia odorifera* | 20 | 2.7 |
|  | *Enterobacter* spp. | 20 | 2.7 |
|  | *Klebsiella* spp. | 20 | 2.7 |
|  | *Salmonella* spp. | 18 | 2.4 |
|  | *Serratia* spp. | 18 | 2.4 |
|  | *Citrobacter freundii* | 16 | 2.1 |
|  | *Proteus* spp. | 11 | 1.5 |
|  | Others* | 58 | 7.8 |
| *Staphylococcus* (n = 367) | Coagulase-positive staphylococci | 228 | 62.1 |
|  | Other staphylococci | 139 | 37.9 |
| *Pseudomonas* (n = 83) | *Pseudomonas aeruginosa* | 48 | 57.8 |
|  | Other *Pseudomonas* spp. | 35 | 42.2 |

\*Others: *Serratia marcescens* (6), *Pantoea agglomerans* (reported as *Enterobacter agglomerans*, 6), *Klebsiella oxytoca* (5), *Citrobacter* spp. (5), *Shigella* spp. (5), *Citrobacter koseri* (5), *Morganella morganii* (5), *Kluyvera* spp. (4), *Providencia* spp. (3), *Yersinia* spp. (3), *Citrobacter youngae* (2), *Klebsiella aerogenes* (2), *Proteus vulgaris* (2), *Pantoea* spp. (2), *Edwardsiella* spp. (1), *Raoultella* spp. (1) and *Citrobacter braakii* (1).

### Agreement between EUCAST 2025 and CLSI 2025

For Enterobacterales, four of the 12 antibiotics tested (33.3%) showed poor agreement between CLSI 2025 and EUCAST 2025: amoxicillin–clavulanic acid (κ = 0.52; concordance 89.4%), cefepime (κ = 0.58; 82.6%), aztreonam (κ = 0.56; 76.6%) and ofloxacin (κ = 0.48; 70.3%) (Table 2).

**Table 2.** Comparison of antimicrobial susceptibility results between EUCAST 2025 and CLSI 2025 among Enterobacterales clinical isolates at Buea Regional Hospital, 2023–2025.

| Antibiotic | Number tested | CLSI | | | EUCAST | | | Concordance (%) | $\kappa$ (95% CI) |
| --- | --- | --- | --- | --- | --- | --- | --- | --- | --- |
|  |  | % S | % I/SDD | % R | % S | % I | % R |  |  |
| Amoxicillin–clavulanic acid | 649 | 7.6 | 9.5 | 82.9 | 6.5 | 0.0 | 93.5 | 89.4 | 0.52 (0.42–0.60) |
| Cefepime | 453 | 17.7 | 12.3 | 70.0 | 12.6 | 6.4 | 81.0 | 82.6 | 0.58 (0.51–0.65) |
| Ceftriaxone | 621 | 19.2 | 4.0 | 76.8 | 12.2 | 6.4 | 81.3 | 88.9 | 0.68 (0.62–0.74) |
| Imipenem | 515 | 73.6 | 11.6 | 14.8 | 78.4 | 7.8 | 13.8 | 94.2 | 0.85 (0.79–0.90) |
| Meropenem | 605 | 71.9 | 13.4 | 14.7 | 77.4 | 11.7 | 10.9 | 90.7 | 0.78 (0.72–0.83) |
| Aztreonam | 552 | 31.2 | 11.8 | 57.1 | 19.6 | 11.6 | 68.8 | 76.6 | 0.56 (0.51–0.61) |
| Gentamicin | 511 | 36.6 | 15.7 | 47.7 | 43.4 | 0.0 | 56.6 | 84.3 | 0.73 (0.68–0.77) |
| Amikacin | 528 | 59.7 | 19.5 | 20.8 | 73.7 | 0.0 | 26.3 | 80.5 | 0.61 (0.56–0.67) |
| Ciprofloxacin | 574 | 20.2 | 12.7 | 67.1 | 23.7 | 9.2 | 67.1 | 96.5 | 0.93 (0.90–0.96) |
| Ofloxacin | 377 | 49.3 | 6.4 | 44.3 | 25.7 | 8.2 | 66.0 | 70.3 | 0.48 (0.42–0.55) |
| Levofloxacin | 533 | 40.9 | 10.5 | 48.6 | 35.8 | 11.1 | 53.1 | 90.4 | 0.84 (0.79–0.88) |
| Trimethoprim–sulfamethoxazole | 485 | 15.3 | 3.7 | 81.0 | 16.5 | 2.5 | 81.0 | 98.8 | 0.96 (0.93–0.99) |

$\kappa < 0.61$ : poor agreement; $\kappa = 0.61–0.80$ : substantial agreement; $\kappa = 0.81–1.00$ : almost perfect agreement. S: susceptible; I: intermediate (CLSI) or susceptible, increased exposure (EUCAST); SDD: susceptible-dose dependent; R: resistant; CI: confidence interval.

For *Staphylococcus* isolates, five (gentamicin, ciprofloxacin, levofloxacin, minocycline and rifampicine) of the 10 antibiotics tested (50%) showed poor agreement between EUCAST 2025 and CLSI 2025 as shown in table 3.

**Table 3.** Comparison of antimicrobial susceptibility results between EUCAST 2025 and CLSI 2025 among *Staphylococcus* clinical isolates at Buea Regional Hospital, 2023–2025.

| Antibiotic | Number tested | CLSI | | | EUCAST | | | Concordance (%) | $\kappa$ (95% CI) |
| --- | --- | --- | --- | --- | --- | --- | --- | --- | --- |
|  |  | % S | % I | % R | % S | % I | % R |  |  |
| Cefoxitin | 356 | 34.8 | 0 | 65.2 | 34.8 | 0 | 65.2 | 100 | 1 |
| Gentamicin | 232 | 74.1 | 6.9 | 19 | 58.2 | 0 | 41.8 | 77.2 | 0.53 (0.45–0.62) |
| Ciprofloxacin | 264 | 42 | 11 | 47 | 0.4 | 50.4 | 49.2 | 56.1 | 0.38 (0.33–0.43) |
| Levofloxacin | 227 | 62.6 | 12.8 | 24.7 | 0.4 | 48.9 | 50.7 | 25.6 | 0.08 (0.03–0.13) |
| Erythromycin | 336 | 26.8 | 11.6 | 61.6 | 31.2 | 0 | 68.8 | 88.4 | 0.76 (0.71–0.82) |
| Clindamycin | 338 | 68.6 | 12.5 | 18.9 | 66 | 0 | 34 | 84.9 | 0.69 (0.63–0.75) |
| Tetracycline | 177 | 46.3 | 6.2 | 47.5 | 41.8 | 0 | 58.2 | 89.3 | 0.79 (0.71–0.88) |
| Minocycline | 194 | 85.1 | 9.8 | 5.1 | 63.9 | 0 | 36.1 | 69.1 | 0.29 (0.21–0.38) |
| Trimethoprim–sulfamethoxazole | 245 | 22.4 | 13.1 | 64.5 | 20 | 7.3 | 72.7 | 89.4 | 0.78 (0.69–0.85) |
| Rifampicin | 257 | 88.7 | 1.9 | 9.3 | 74.3 | 0 | 25.7 | 83.7 | 0.48 (0.36–0.60) |

$\kappa < 0.61$ : poor agreement; $\kappa = 0.61–0.80$ : substantial agreement; $\kappa = 0.81–1.00$ : almost perfect agreement. S: susceptible; I: intermediate (CLSI) or susceptible, increased exposure (EUCAST); R: resistant; CI: confidence interval.

For *Pseudomonas* isolates, five (cefepime [κ = 0.24; 46%], imipenem [κ = 0.24; 35.8%], aztreonam [κ = 0.37; 54.4%], ciprofloxacin [κ = 0.29; 56.7%] and levofloxacin [κ = 0.38; 53.8%]) of the seven antibiotics tested (71.4%) showed poor agreement between EUCAST 2025 and CLSI 2025 as shown in table 4.

**Table 4.** Comparison of antimicrobial susceptibility results between EUCAST 2025 and CLSI 2025 among *Pseudomonas* isolates at Buea Regional Hospital, 2023–2025.

| Antibiotic | Number tested | CLSI | | | EUCAST | | | Concordance (%) | $\kappa$ (95% CI) |
| --- | --- | --- | --- | --- | --- | --- | --- | --- | --- |
|  |  | % S | % I | % R | % S | % I | % R |  |  |
| Cefepime | 50 | 54 | 2 | 44 | 0 | 36 | 64 | 46 | 0.24 (0.16–0.33) |
| Imipenem | 67 | 65.7 | 3 | 31.3 | 4.5 | 61.2 | 34.3 | 35.8 | 0.24 (0.15–0.32) |
| Meropenem | 66 | 56.1 | 3 | 40.9 | 51.5 | 13.6 | 34.8 | 89.4 | 0.81 (0.69–0.92) |
| Aztreonam | 57 | 42.1 | 19.3 | 38.6 | 0 | 57.9 | 42.1 | 54.4 | 0.37 (0.24–0.49) |
| Amikacin | 61 | 72.1 | 3.3 | 24.6 | 75.4 | 0 | 24.6 | 96.7 | 0.92 (0.80–1) |
| Ciprofloxacin | 67 | 35.8 | 10.4 | 53.7 | 1.5 | 32.8 | 65.7 | 56.7 | 0.29 (0.19–0.39) |
| Levofloxacin | 52 | 50 | 17.3 | 32.7 | 7.7 | 55.8 | 36.5 | 53.8 | 0.38 (0.23–0.52) |

$\kappa < 0.61$ : poor agreement; $\kappa = 0.61–0.80$ : substantial agreement; $\kappa = 0.81–1.00$ : almost perfect agreement. S: susceptible; I: intermediate (CLSI) or susceptible, increased exposure (EUCAST); R: resistant; CI: confidence interval.

Fig 1 compares the susceptibility percentages of Enterobacterales isolates obtained using EUCAST 2025 and CLSI 2025 breakpoint interpretations. The surveillance impact varied across the antibiotics tested. Remarkable differences were observed for ofloxacin, amikacin and aztreonam. Ofloxacin showed the widest discrepancy, with susceptibility decreasing remarkably under EUCAST compared with CLSI.

**Fig 1.**
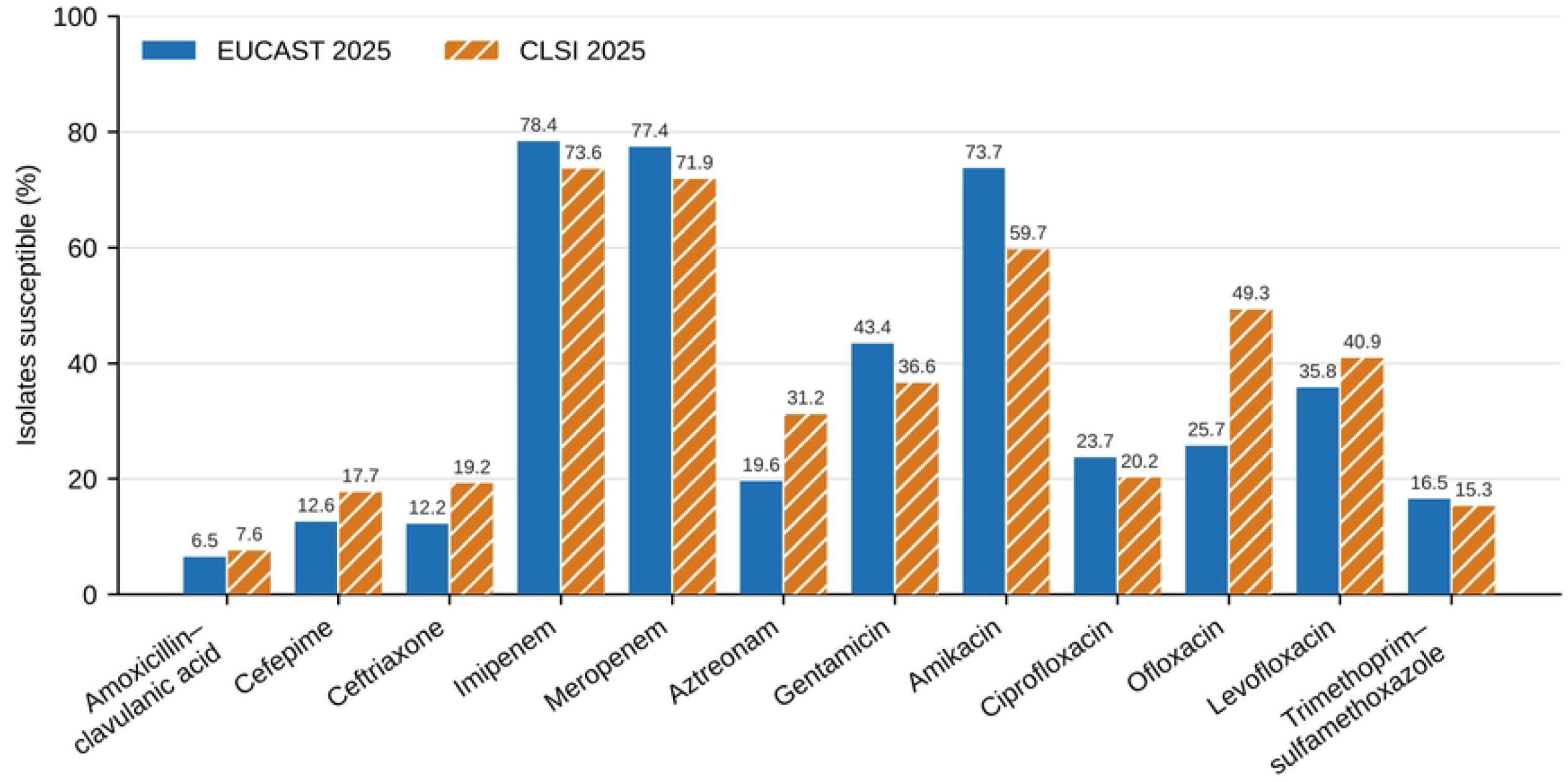
Antibiogram comparison between EUCAST 2025 and CLSI 2025 among Enterobacterales clinical isolates at Buea Regional Hospital, 2023–2025.

When comparing the susceptibility percentages of *Staphylococcus* isolates under EUCAST 2025 and CLSI 2025, ciprofloxacin and levofloxacin showed the widest discrepancies, with susceptibility evidently lower under EUCAST than CLSI. Gentamicin, minocycline and rifampicin also showed remarkable differences in susceptibility percentages (Fig 2).

**Fig 2.**
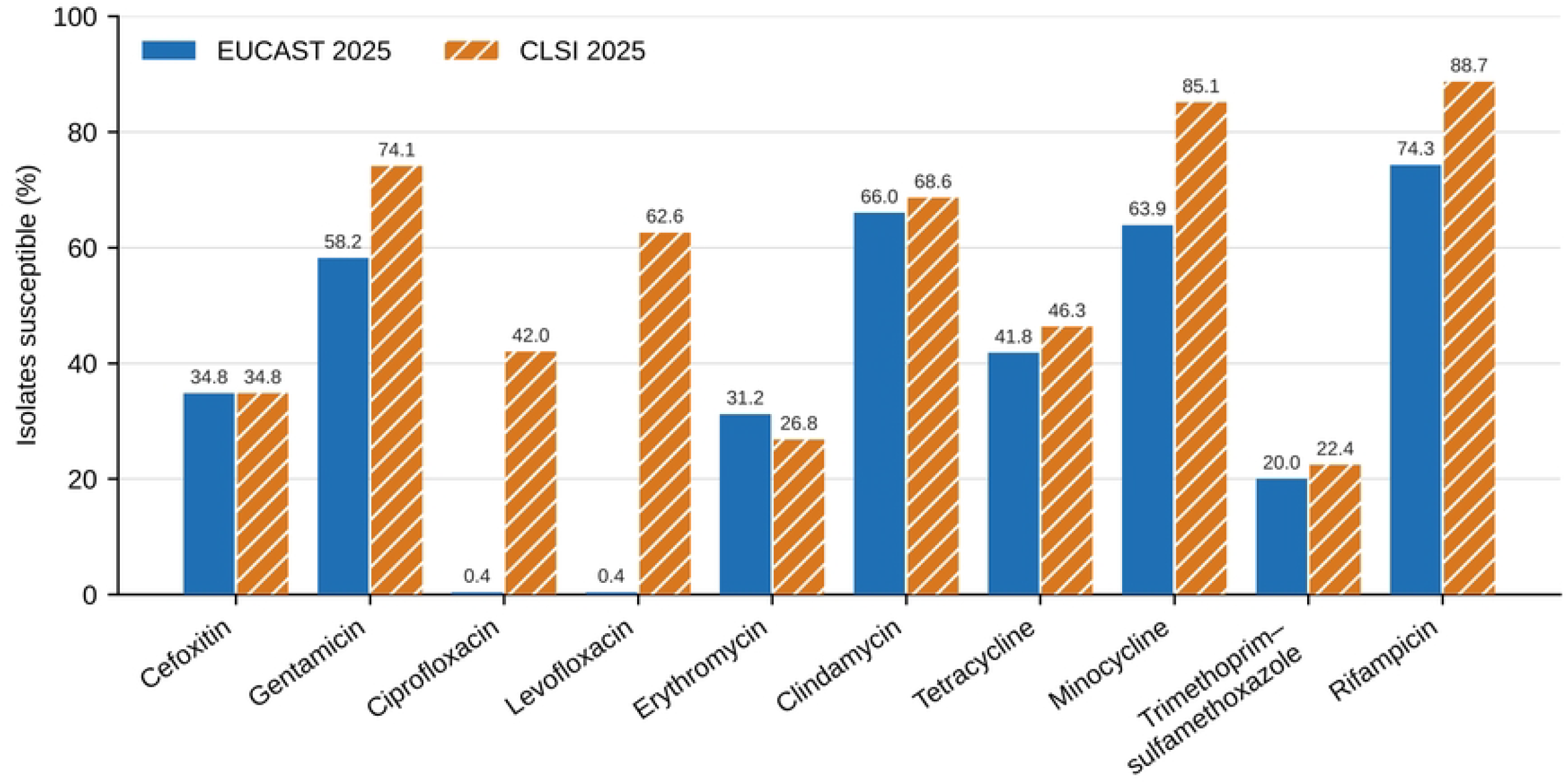
Antibiogram comparison between EUCAST 2025 and CLSI 2025 among *Staphylococcus* clinical isolates at Buea Regional Hospital, 2023–2025.

For the comparison of susceptibility percentages of *Pseudomonas* isolates, CLSI classified more isolates as susceptible than EUCAST for all antibiotics tested except amikacin (Fig 3).

**Fig 3.**
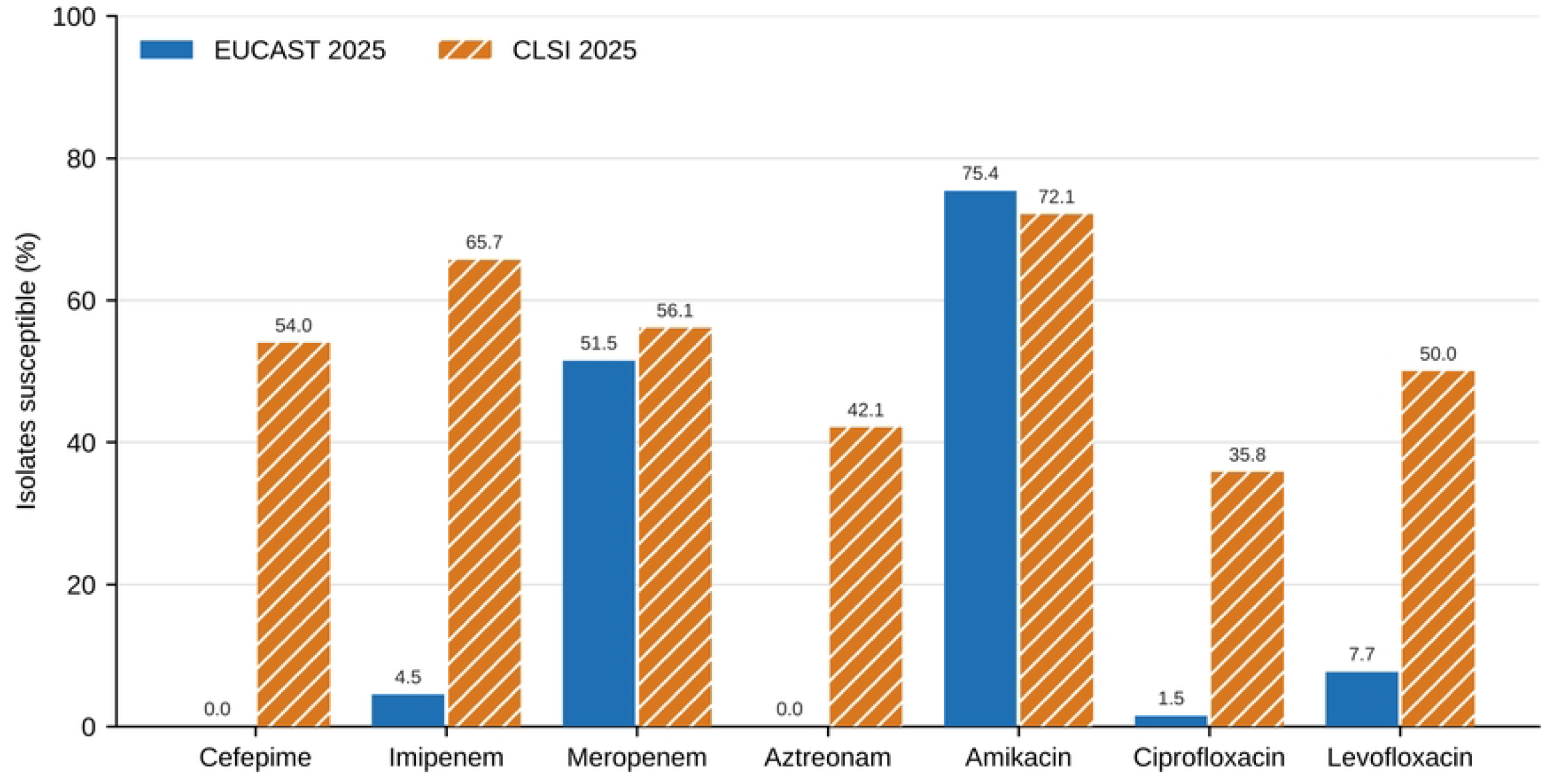
Antibiogram comparison between EUCAST 2025 and CLSI 2025 among *Pseudomonas* clinical isolates at Buea Regional Hospital, 2023–2025. Major interpretation switches (S ↔ R)

For Enterobacterales, three (amoxicillin–clavulanic acid [7/649 isolates, 1.1%], ceftriaxone [4/621 isolates, 0.6%] and ofloxacin [59/377 isolates, 15.6%]) of the 12 antibiotics tested recorded major interpretation switches (S ↔ R) between CLSI 2025 and EUCAST 2025: For *Staphylococcus* isolates, six (gentamicin [37/232 isolates, 15.9%], levofloxacin [31/227, 13.7%], clindamycin [9/338, 2.7%], tetracycline [8/177, 4.5%], minocycline [41/194, 21.1%] and rifampicin [37/257, 14.4%] of the 10 antibiotics tested recorded major interpretation switches as shown in table 5.

**Table 5.** Frequency and percentage of major interpretation switches (S ↔ R) between EUCAST 2025 and CLSI 2025 among Enterobacterales, *Staphylococcus* and *Pseudomonas* isolates at Buea Regional Hospital, 2023–2025.

| Pathogen group | Antibiotic | Number tested | Number of major switches (S ↔ R) | Major switches (%) |
| --- | --- | --- | --- | --- |
| Enterobacterales | Amoxicillin–clavulanic acid | 649 | 7 | 1.1 |
|  | Cefepime | 453 | 0 | 0 |
|  | Ceftriaxone | 621 | 4 | 0.6 |
|  | Imipenem | 515 | 0 | 0 |
|  | Meropenem | 605 | 0 | 0 |
|  | Aztreonam | 552 | 0 | 0 |
|  | Gentamicin | 511 | 0 | 0 |
|  | Amikacin | 528 | 0 | 0 |
|  | Ciprofloxacin | 574 | 0 | 0 |
|  | Ofloxacin | 377 | 59 | 15.6 |
|  | Levofloxacin | 533 | 0 | 0 |
|  | Trimethoprim–sulfamethoxazole | 485 | 0 | 0 |
| *Staphylococcus* | Cefoxitin | 356 | 0 | 0 |
|  | Gentamicin | 232 | 37 | 15.9 |
|  | Ciprofloxacin | 264 | 0 | 0 |
|  | Levofloxacin | 227 | 31 | 13.7 |
|  | Erythromycin | 336 | 0 | 0 |
|  | Clindamycin | 338 | 9 | 2.7 |
|  | Tetracycline | 177 | 8 | 4.5 |
|  | Minocycline | 194 | 41 | 21.1 |
|  | Trimethoprim–sulfamethoxazole | 245 | 0 | 0 |
|  | Rifampicin | 257 | 37 | 14.4 |
| *Pseudomonas* | Cefepime | 50 | 10 | 20 |
|  | Imipenem | 67 | 0 | 0 |
|  | Meropenem | 66 | 0 | 0 |
|  | Aztreonam | 57 | 0 | 0 |
|  | Amikacin | 61 | 0 | 0 |
|  | Ciprofloxacin | 67 | 2 | 3 |
|  | Levofloxacin | 52 | 0 | 0 |

S: susceptible; R: resistant.

### Breakpoint system more likely to classify isolates as non-susceptible

Generally, EUCAST 2025 was more likely than CLSI 2025 to classify Enterobacterales, *Staphylococcus* and *Pseudomonas* isolates as non-susceptible, with a statistically significant difference (p < 0.001) (Table 6).

**Table 6.** Comparison of non-susceptibility classification rates between EUCAST 2025 and CLSI 2025 among Enterobacterales, *Staphylococcus* and *Pseudomonas* isolates at Buea Regional Hospital, 2023–2025.

| Organism group | Results classified as non-susceptible by EUCAST but susceptible by CLSI | Results classified as susceptible by EUCAST but non-susceptible by CLSI | Exact McNemar p-value | Guideline more likely to classify isolates as non-susceptible |
| --- | --- | --- | --- | --- |
| Enterobacterales | 253 | 193 | 0.005 | EUCAST |
| *Staphylococcus* spp. | 389 | 15 | $< 0.001$ | EUCAST |
| *Pseudomonas* spp. | 140 | 2 | $< 0.001$ | EUCAST |
| Overall | 782 | 210 | $< 0.001$ | EUCAST |

Non-susceptible = I, SDD or R.

## Discussion

This study demonstrates that EUCAST 2025 and CLSI 2025 do not always produce the same AST interpretation when applied to the same inhibition-zone measurements. Such differences probably arise because the two standards may use different zone-diameter breakpoints, definitions of susceptibility categories and dosing assumptions. Other studies have reported similar changes in susceptibility classification and cumulative antibiograms when laboratories moved from one standard to the other [7,8]. In the present study, the proportion of antibiotics showing poor agreement increased from 33.3% among Enterobacterales to 50.0% among *Staphylococcus* spp. and 71.4% among *Pseudomonas* spp. This shows that the two guideline systems cannot be viewed as completely interchangeable and that the consequence of guideline selection varies according to the organism–antibiotic combination. Understanding the reasons for the disagreement therefore requires examination of the individual antibiotics affected within each bacterial group.

Among Enterobacterales, poor agreement involved amoxicillin–clavulanic acid, cefepime, aztreonam and ofloxacin. For cefepime, much of the disagreement reflects how each standard handles isolates that may still respond to higher doses: CLSI labels them SDD, whereas EUCAST labels them “I”, and the zone diameter ranges assigned to these categories do not coincide. Ofloxacin showed the lowest agreement, while ciprofloxacin and levofloxacin agreed closely. EUCAST and CLSI have similar break points for Levofloxacin and Ciprofloxacin. For ofloxacin, CLSI considers susceptibility when zone diameter is ≥16mm while EUCAST considers susceptibility as from 24mm. The problem is therefore specific to the ofloxacin breakpoints rather than to fluoroquinolones as a class. For amoxicillin– clavulanic acid, CLSI classified more isolates as susceptible, consistent with a Belgian study in which CLSI categorized more *E. coli* isolates as susceptible as compared to EUCAST [17]. The aminoglycosides performed otherwise, with EUCAST reporting a higher proportion of susceptible isolates than CLSI for gentamicin and amikacin. A plausible justification for this could be the fact that EUCAST applies no intermediate category to these agents, whereas the CLSI does. Poor agreement between the standards is therefore bidirectional, even though there is an overall non-susceptibility favoritism with EUCAST. A more thrilling form of this breakpoint-placement effect occurred among the Gram-positive isolates.

Half (50%) of the antibiotics tested against *Staphylococcus*, showed poor agreement with levofloxacin and ciprofloxacin recording the poorest outcome. CLSI susceptibility cut-off for levofloxacin and ciprofloxacin is 19 mm and 21mm respectively while EUCAST considers susceptibility as from 50mm for both antibiotics, a zone size that many Staphylococcal isolates practically never attain. Looking at the wide breakpoint gap, it is evident that many Staphylococci that are susceptible to these antibiotics as per CLSI would be considered non-susceptible with EUCAST. That explains why in this study, the proportion of staphylococcal isolates reported as susceptible fell from 42% to 0.4% for ciprofloxacin and from 62.6% to 0.4% for levofloxacin when EUCAST was applied. Hearteningly, cefoxitin interpretations were identical under both standards, so the detection of methicillin resistance, which underpins both MRSA infection control and MRSA surveillance, was unaffected by the choice of standard. The same exposure-based logic, applied to an organism with intrinsically limited antibiotic permeability, produced the widest divergence of all.

*Pseudomonas* was the group most affected by the choice of standard with 71.4 % of antibiotics tested revealing poor agreement. EUCAST considers the same 50mm off-scale susceptible breakpoint approach to cefepime, aztreonam, imipenem, ciprofloxacin and levofloxacin for *Pseudomonas*, reiterating its stands that these organisms maybe susceptible to these agents only at high doses. As such, zone diameters reported as susceptible under CLSI were considered as “I” under EUCAST. For example, under CLSI, 54% of isolates were susceptible to cefepime while none were susceptible to the same antibiotic under EUCAST. This divergence is clinically relevant because β-lactams remain central to the treatment of *P. aeruginosa* infections [18]. A laboratory using EUCAST would report that only high-dose regimens are effective, whereas one using CLSI would report many of the same *Pseudomonas* isolates as susceptible at standard doses. Disagreement that merely moves isolates into or out of an increased-exposure category (I or SDD) can usually be handled by dose adjustment. The main concern arises when one standard says “susceptible” and the other outrightly says “resistant” for the same zone diameter on the same isolate.

Major interpretation switches are the most significant form of discordance, because they reverse the verdict on whether an antibiotic can be used at all in a given situation. Major interpretation switches were recorded in three of 12 antibiotics (25%) among Enterobacterales, six of 10 (60%) among *Staphylococcus* and two of seven (28.6%) among *Pseudomonas*. Switches (irrespective of the direction) may come with serious consequences. A false susceptible result may lead to therapeutic failure if the clinician decides to prescribe the antibiotic in question, whereas a false resistant result may prompt the clinician to consider more expensive, broad spectrum, reserved antibiotics like carbapenems there by increasing costs and selection pressure. Beyond the individual patient, these reclassifications and associated consequences extend into every dataset built from routine results.

The surveillance implications are particularly relevant for Cameroon because laboratories using either standard donate data to the same national dataset periodically. Our findings reveal how large the resulting bias can be: for example, levofloxacin susceptibility was 62.6% under CLSI but 0.4% under EUCAST for *Staphylococcus*. Differences in AMR between surveillance sites may therefore reflect their choice of standards used rather than true geographical variation, which could misguide national antibiograms and stewardship priorities. However, because WHONET stores raw zone diameters, one way of minimizing this problem could be the reinterpretation of all surveillance site data once all is received and merged at the central level using a single standard. The big question now is, which of the two standards should be prioritized for the harmonized data and why?

EUCAST classified significantly more isolates as non-susceptible than CLSI in all three groups. This is consistent with the findings of other studies [4,7]. This implies EUCAST may prompt earlier dose optimization and reduce the risk of therapeutic failure especially in severe infections, but on the downside, it can push clinicians towards reserve antibiotics even in cases where watch or access antibiotics could have been effective. On the other hand, CLSI optimizes the use of cheaper, narrower-spectrum antibiotic options for patients, but risks tagging as susceptible some isolates that standard antibiotic doses will not reliably kill or inhibit their growth in vivo. Neither standard possesses monopoly power over biological “truth” in every case. So which standard should therefore be prioritized? The choice depends on which type of error the country (or hospital) in question is willing to tolerate in its setting: false resistance or false susceptibility. Another important factor to take into consideration is cost and sustainability: EUCAST breakpoint tables are freely available online meanwhile CLSI requires payment before access. Both standards go through yearly updates and so loyal users must be committed to acquiring the most recent version yearly. This implies CLSI users must be capable of affording the new versions yearly if not, sustainability becomes questionable.

This study had some limitations. Firstly, the comparison was based only on disk diffusion method and zone diameters close to breakpoints are liable to measurement inconsistency. MIC outcome may be different and cannot be overemphasized. Secondly, the study compared laboratory interpretations without clinical outcome data, so it could not determine which standard better forecasts therapeutic success. Finally, the study was conducted in a single hospital laboratory and so cannot be generalized. The extent of discordance/concordance may differ at other surveillance sites.

In conclusion, poor agreement between EUCAST 2025 and CLSI 2025 affected one third of the antibiotics tested for Enterobacterales, half for *Staphylococcus* and close to three quarters for *Pseudomonas*. Major interpretation switches were quite common for levofloxacin, ofloxacin, gentamicin, minocycline, rifampicin and cefepime. EUCAST was unswervingly and significantly more likely to classify isolates as non-susceptible. The national AMR surveillance program in Cameroon should consider adopting a single breakpoint standard for all its surveillance sites so as to guarantee uniformity and optimize the reliability of its surveillance data.

## Data Availability

All relevant data are within the manuscript and its Supporting information files.

## Acknowledgments

Special thanks to the management and laboratory personnel of Buea Regional Hospital for permission to conduct this study and for extracting and de-identifying the laboratory records.

## Supporting information

**S1 File. De-identified isolate-level dataset with raw inhibition zone diameters and EUCAST 2025 and CLSI 2025 interpretations, Buea Regional Hospital, 2023–2025**. (XLSX)

